# Epidemiological Transition of Covid-19 in India from Higher to Lower HDI States and Territories: Implications for Prevention and Control

**DOI:** 10.1101/2020.05.05.20092593

**Authors:** Rajeev Gupta, Rajinder K Dhamija, Kiran Gaur

**Author notes:** Correspondence:* Dr Rajeev Gupta, Department of Medicine, Eternal Heart Care Centre & Research Institute, Jagatpura Road, Jawahar Circle, Jaipur 302017 India.

## Abstract

**Background & Objective:** Social determinants of evolving covid-19 pandemic have not been well studied. To determine trends in transition of this epidemic in India we performed a study in states at various levels of human development index (HDI).

**Methods:** We used publicly available data sources to track progress of covid-19 epidemic in India in different states and territories where it was reported in significant numbers. The states (n=20) were classified into tertiles of HDI and weekly trends in cases and deaths plotted from 15 March to 2 May 2020. To assess association of HDI with state-level covid-19 burden we performed Pearson’s correlation. Logarithmic trends were evaluated for calculation of projections. A microlevel study was performed in select urban agglomerations for identification of socioeconomic status (SES) differentials.

**Results:** There is wide regional variation in covid-19 cases and deaths in India from mid-March to early-May 2020. High absolute numbers have been reported from states of Maharashtra, Gujarat, Delhi, Madhya Pradesh, Rajasthan and Tamilnadu. Growth rate in cases and deaths is slow in high HDI states while it has increased rapidly in middle and lower HDI states. In mid-March 2020 there was a strong positive correlation of state-level HDI with weekly covid-19 cases (r= 0.37, 0.40) as well as deaths (r= 0.31, 0.42). This declined by early-May for cases (r= 0.04, 0.06) as well as deaths (r= - 0.005, 0.001) with significant negative logarithmic trend (cases R squared= 0.92; deaths R squared= 0. 84). These trends indicate increasing cases and deaths in low HDI states. Projection reveals that this trend is likely to continue to early-June 2020. Microlevel evaluation shows that urban agglomerations are major focus of the disease in India and it has transited from middle SES to low SES locations.

**Conclusion:** There is wide variability in burden of covid-19 in India. Slow growth and flattening of curve is observed in high-HDI states while disease is increasing in mid and lower HDI states. Projections reveal that lower HDI states would achieve parity with high HDI states by early-June 2020. Covid-19 is mostly present in urban agglomerations where it has transited from upper-middle to low SES locations. Public health strategies focusing on urban low SES locations and low HDI states are crucial to decrease covid-19 burden in India.

## INTRODUCTION

As India enters the eighth week of the successful nationwide lockdown due to pandemic of novel coronavirus related severe acute respiratory syndrome (SARS CoV-2, Covid-19) there is a need to focus on social and public health science. This is important for the life to reboot, for students to go back to schools and colleges, offices to function, factories to commence production and governance to step-up. Without science, we cannot resume life and it is important for policy makers and health bureaucrats to plan further epidemic control strategies.

An important rule of epidemiological transition of a particular disease is that it starts from the rich provinces, districts, locations and high socioeconomic status (SES) individuals and then gradually permeates down to middle SES and finally comes to rest among the low SES people in low SES locations.^1^ This trend has not only been visible for infectious diseases but also for non-communicable diseases across the globe.^2,3^ Numerous examples exist.^4^ Plague from the Justinian era to the late nineteenth century, cholera epidemics from time immemorial to middle of the last century, spread of malaria and yellow-fever from Africa and Asia to Europe and north America, influenza-A pandemic exactly a hundred years ago and the current coronavirus related SARS have all followed this path of transition from rich to the poor along the lines of commerce and business. The first plague pandemic (Justinian plague) in 6^th^-7^th^ centuries is lost to memories but secular track of the *Black Death* in fourteenth century Europe is well known. The epidemic began in central and eastern Asia in 1340’s, travelled along the caravan-routes with businessmen and initially infected upper SES people in central and southern Europe, western Asia and northern Africa before spreading in the poor colonies of the major cities along the trade routes. When sea-faring started in earnest and ships became the conduits of business, plague travelled along these ships from eastern, southern and western Asian ports among business people and administrators and quickly spread to the poor working in ships and their families in coastal cities before moving inland among the low SES. Similar track was mapped for cholera, which was carried from southern and eastern Asian ports among businessmen and seamen to North Africa, Europe and North America and rapidly spread among the poor men, women and children in port cities and elsewhere. Yellow fever also spread from shipmen and slaves from Africa to business people in Europe and the Americas and then to the poor.^4^

Influenza-A epidemic spread from South Africa and Europe along with veterans of the 1^st^ World War and spread rapidly among general population of Europe, Asia and northern Africa and finally landed in Asia where it killed more poor people than the rich in India.^5^ The current covid-19 epidemic began from Wuhan, an industrial city in south-eastern China,^6,7^ and rapidly spread via business-men to populations in Europe, West Asia and North America.^8^ The infection initially was among the high SES people living in upper and mid SES in major cities (Amsterdam, Lombardia, London, Madrid, New York, Tehran, etc.)^9-14^ before spreading to the low SES immigrants and others living in poorer locations in some of these cities.^15^ These transitions indicate that for the epidemic to evolve, mature and stabilize it is essential to understand the dynamics at both macro- and microlevel. We performed a study in India to understand the dynamics of covid-19 epidemic at the macrolevel by examining association of state-level development with time-trends in number of cases and deaths. We have supplemented these macrolevel associations with microlevel case-studies to understand the progress of this epidemic.

## METHODS

The present study has been conducted using publicly available data from multiple sources.

No ethics clearances were obtained as this involved use of secondary data. Daily data on covid-19 in various states and regions of India were obtained from a non-commercial public website www.covid19india.org. This website updates daily data on covid-19 related cases, deaths, recovery and testing at state and district level of the country.^16^ We obtained data only for states (n=20) with a significant number of cases (≥40 cases).

The data on daily cases beginning 15^th^ March 2020, when the cases were beginning to be reported in India, to the present were collated on MS Excel spreadsheets. Data regarding cumulative numbers at weekly intervals for all the states where data were available were calculated. We plotted data on cumulative cases and deaths in each state with significant number of patients using the MS Power Point graphics software. Log transformation was performed for graphs. We obtained data on Human Development Index (HDI) for each state in the country from World Bank site (Supplementary Table 1).^17^ To determine association of state-level HDI with covid-19 cases and deaths we calculated Pearson’s correlation coefficient (r value) at weekly time point. The time-trend related r-values were plotted in a graph using MS Excel. Logarithmic trendline was drawn on the graph to estimate trends (R^2^) and to forecast the correlation at 1 month following the last available data point on 2 May 2020.

## RESULTS

Weekly data on covid-19 cases and deaths in various states of the country is shown in Table 1.^16^ The disease related cases were low in early March 2020 but increased rapidly with time. Significant diversity is observed in absolute and cumulative number of cases and deaths across various states of the country. High absolute number of cases (>2500) is observed in Maharashtra, Gujarat, Delhi, Madhya Pradesh, Rajasthan and Tamilnadu. High rate of increase in cases (logarithmic R^2^ value >0.95) is observed in Haryana, Karnataka, Kerala and Telangana while low rate of increase is not observed in any. High absolute number of covid-19 related deaths is observed in Gujarat and Maharashtra (Table 1).

**Table 1:**
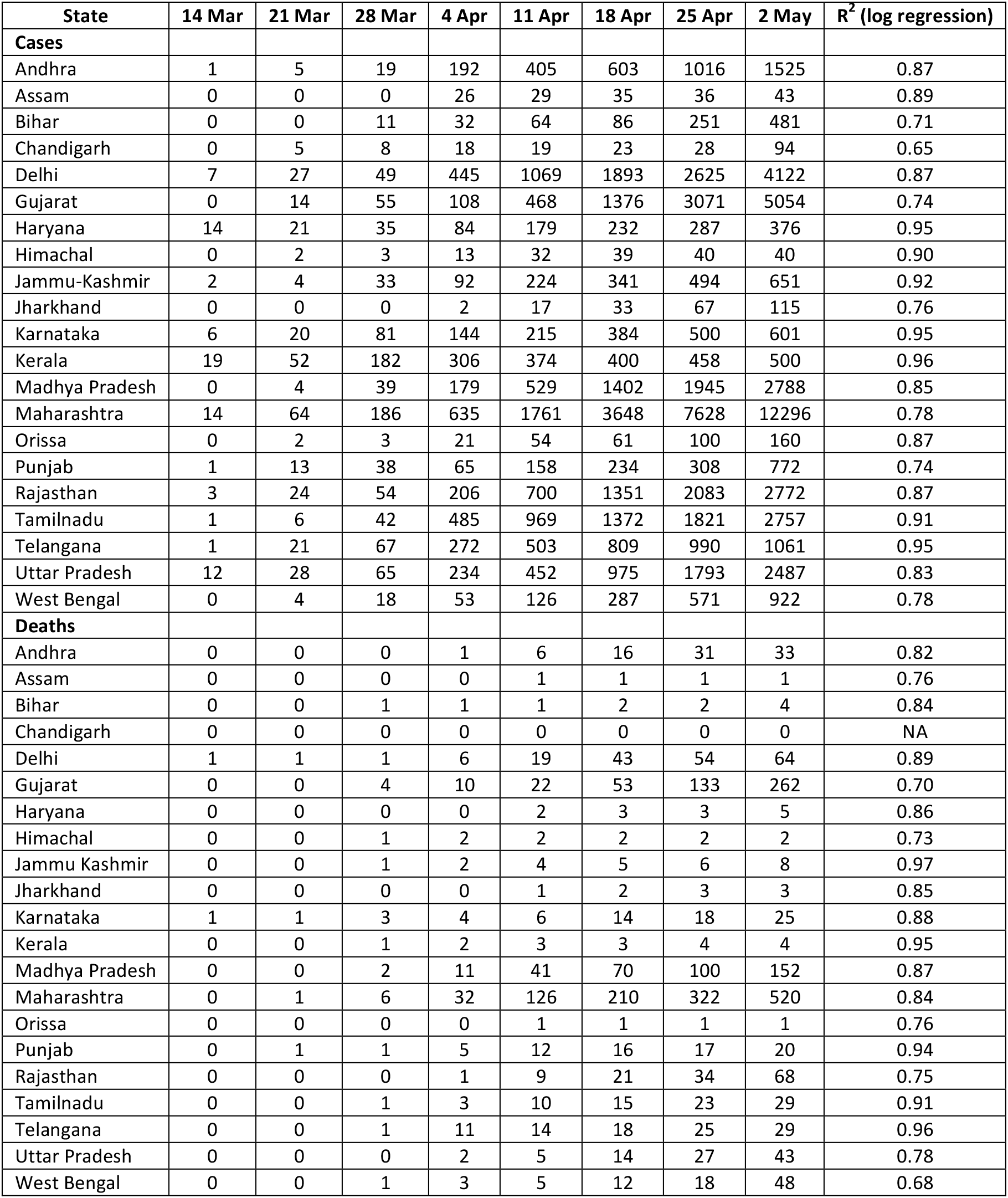
Weekly cumulative number of cases and deaths in different states of the India (alphabetic) in the Months of March and April 2020.

Time trends in cumulative number of confirmed cases of covid-19 in different states of the country are shown in Figure 1. Logarithmic trends of weekly numbers are plotted. Most states in the upper HDI tertile show flattening of the curve, especially Kerala and Himachal. In the mid tertile there is increase in a few states (Maharashtra, Gujarat) while others have reported a gradual ascent. In low tertile group of states there is an ncrease in most states with greatest increase in Rajasthan and Madhya Pradesh. Time trends in cumulative number of deaths in each state in shown in Figure 2. Similar to number of cases, there are large differences in logarithmic trajectory of increase in covid-19 related deaths in different Indian states. Slow increase with flattening of curve is observed in most upper HDI states especially Kerala, Tamilnadu and Himachal. In mid-HDI tertile states, large and rapid increase is observed in Maharashtra, Gujarat and West Bengal while in lower HDI tertile states almost all states demonstrate a rapid increase with high rates in Madhya Pradesh and Rajasthan.

**Figure 1:**
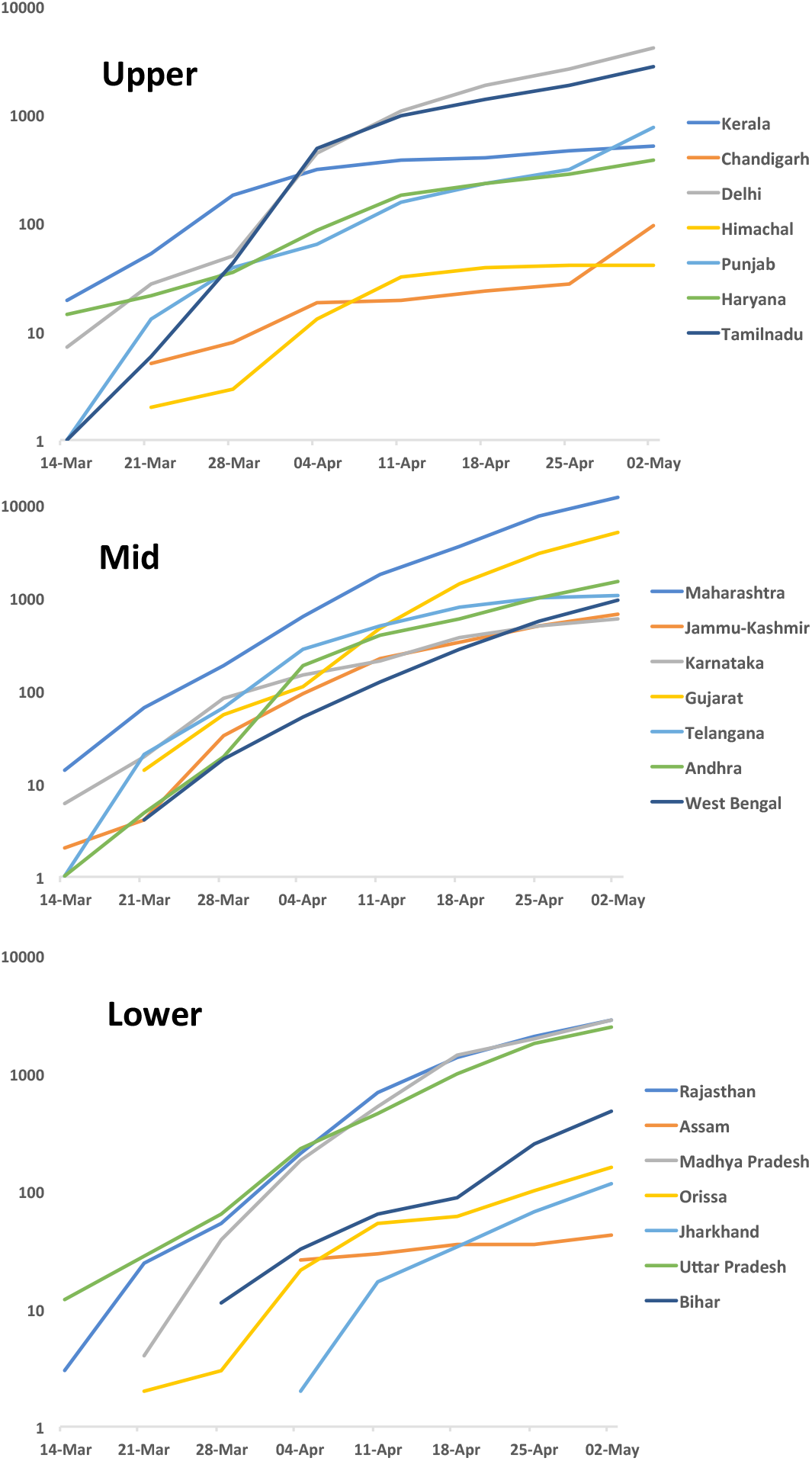
Logarithmic trends in weekly covid-19 related cases (cumulative numbers) in different states and territories of India based on HDI tertiles

**Figure 2:**
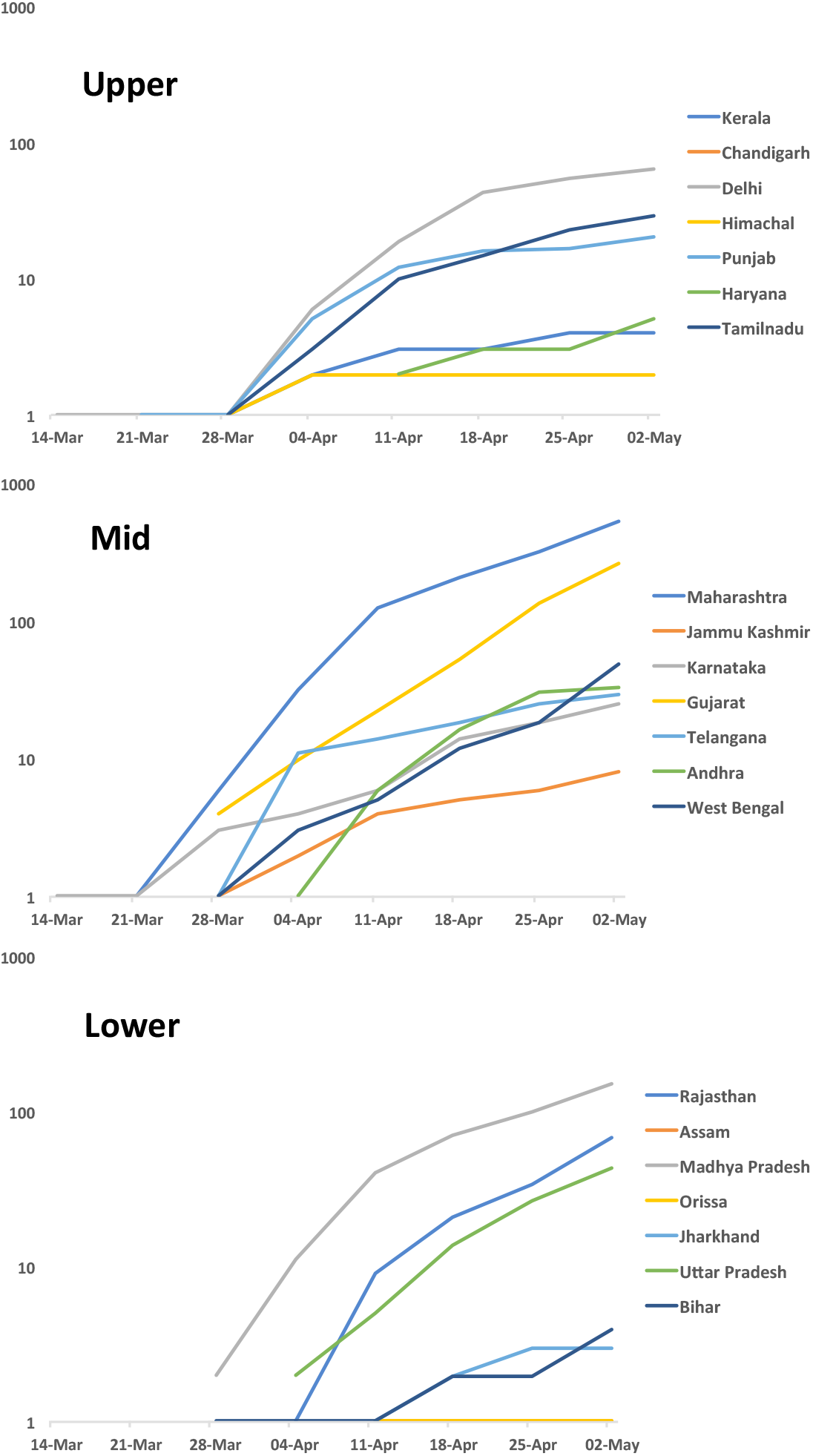
Logarithmic trends in weekly covid-19 related deaths in different states and territories in India based on HDI tertiles

We correlated total covid-19 cases and associated deaths at weekly intervals with HDI of different states to determine their association with development of each state using Pearson’s correlation (Figure 3). In the first two weeks of study there was a strong positive correlation of HDI with covid-19 cases (r= 0.37, 0.40) as well as deaths (r= 0.31, 0.42). This indicates that the disease was more common in better-developed states in March 2020. However, there is a gradual reduction in strength of association and towards early May 2020 a neutral or negative correlation is observed both for cases (r= 0.04, 0.06) as well as deaths (r= −0.005, 0.001) indicating the disease is trending towards lower developed states (Figure 3). A significant negative logarithmic trend is observed both for cases (y= −377.9ln(x)+4040.2, R^2^= 0.92) as well as deaths (y= −351.2ln(x)+3754.9, R^2^= 0.84). Projection of the logarithmic regression estimation suggests that the trend of negative correlation of HDI with covid-19 is likely to continue to end-May or early-June 2020. A significant negative correlation indicates that by this time the epidemic shall be significantly greater in states with lower HDI at this time.

**Figure 3:**
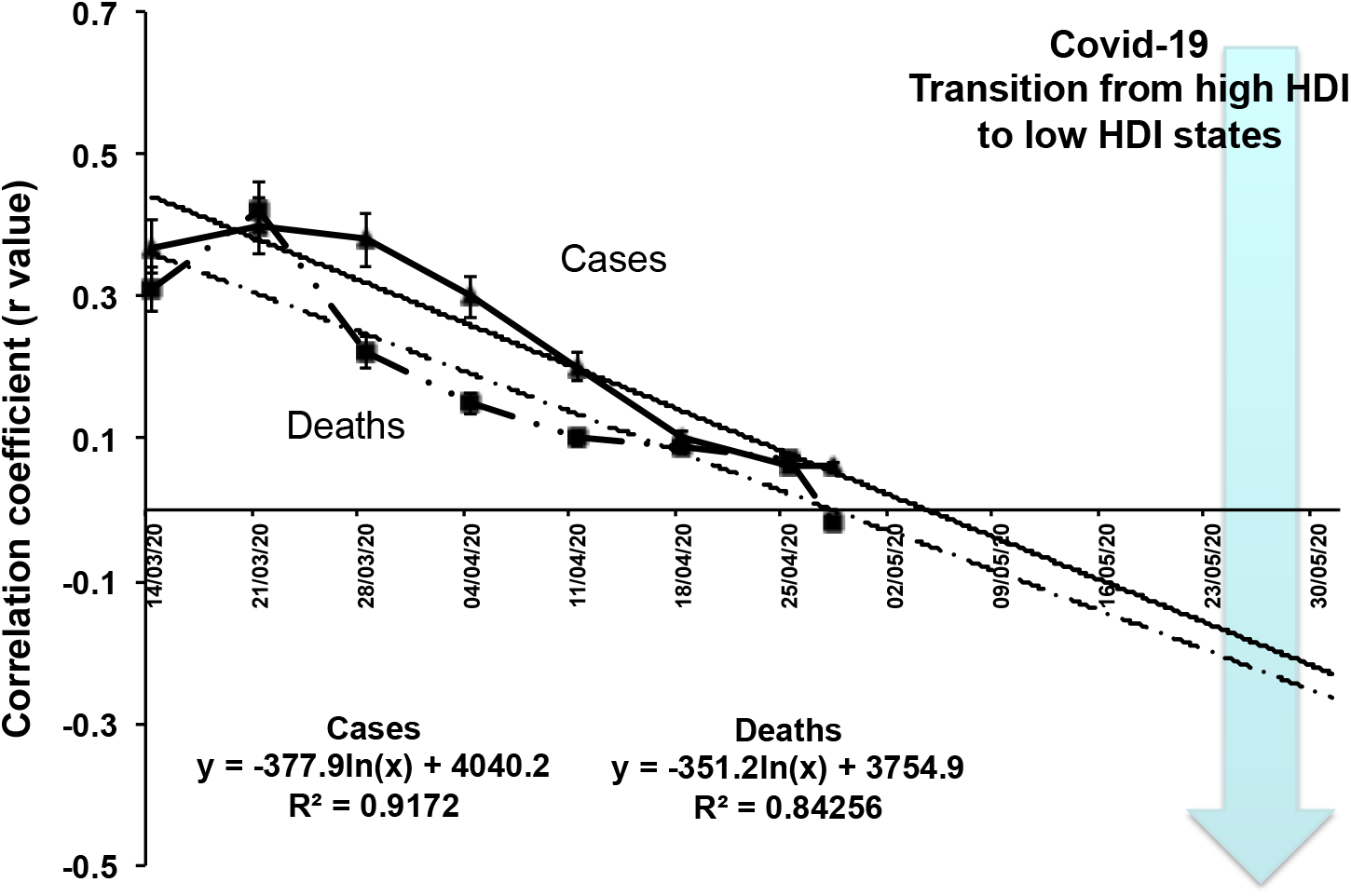
Correlation of state level human development index with weekly cumulative number of cases (solid lines) and deaths (dotted lines) due to Covid-19. Straight line indicate log-regression trends with projection to 30 May 2020.

It has been reported that about 50% of covid-19 cases in India are from 6 urban agglomerations (Mumbai 8359, Delhi 4122, Ahmedabad 3543, Indore 1550, Pune 1339 and Jaipur 961; data on 2^nd^ May 2020).^16^ To determine trends in location-based covid-19 cases and deaths in cities of Jaipur, Delhi and Mumbai we performed microlevel evaluation. In Jaipur (population 2.7 million), the disease was initially reported in travellers, business-persons or students travelling from Italy, Gulf Countries or US in late February to early March 2020, similar to other initial cases in India.^18^ These professionals and business people were living in upper SES locations. Within two weeks rapidly increasing number of patients were reported in multiple areas and presently the highest concentration is in low SES locations in the city (Ramganj 521, Ghatgate 52, Shasrtri Nagar 35 and Johari Bazar 29) (Figure 4). Within these hot spots most of the patients are from low SES. In Mumbai (population 12.6 million) also the disease and deaths were initially reported from up-market locations in the city suburbs, however presently the hotspots are urban slums (Dharavi, Worli Koliwada, etc) (Figure 4).^19^ Similar situation exists in Delhi (population 10.9 million) where the disease has rapidly spread from the foreign-returned travellers and students to low SES locations in various districts of the city as marked in Figure 4.^19^

**Figure 4:**
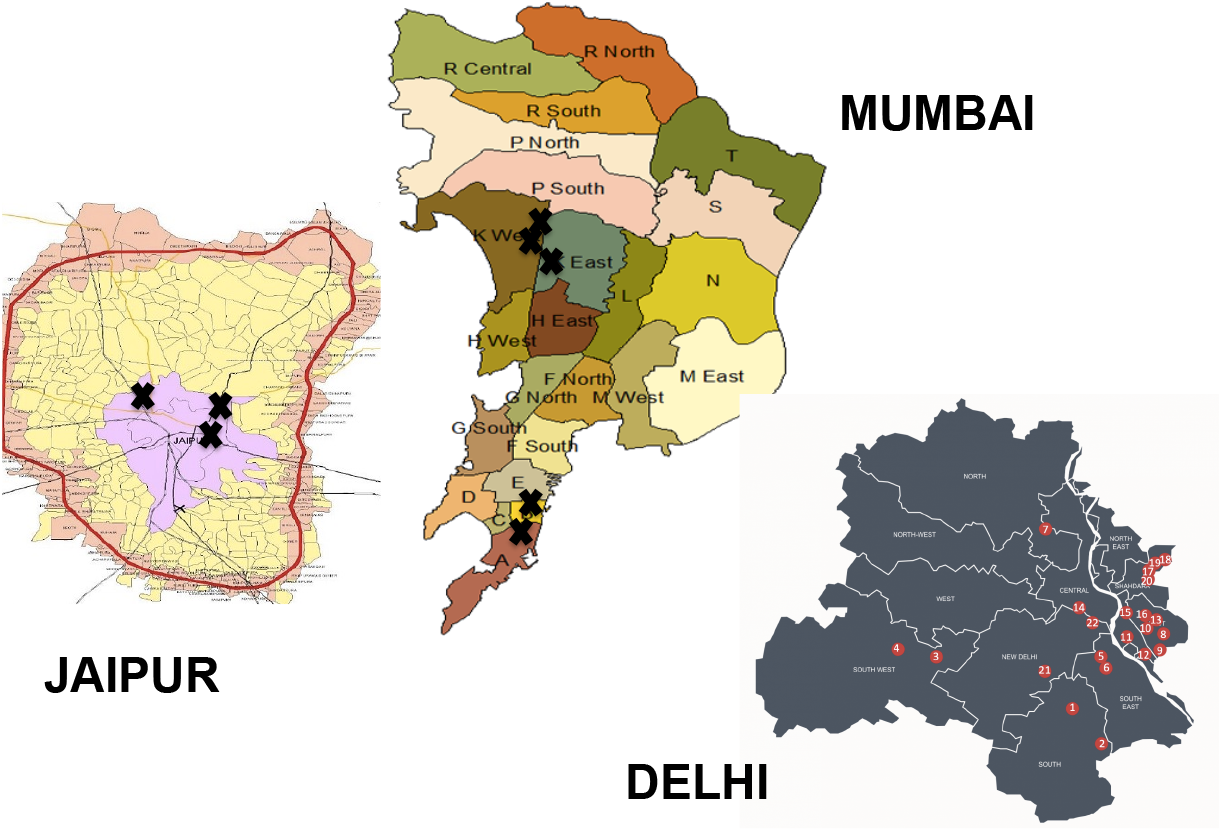
Hotspots of covid-19 cases and deaths in 3 important urban agglomerates in India-Jaipur (left), Mumbai (centre) and Delhi (right). The marked areas are current covid-19 hotspots.

## DISCUSSION

Our analysis shows that covid-19 epidemic in India is following the classical path of epidemiological transition and is still evolving.^2^ Covid-19 related disease and deaths were initially observed in high HDI states. Secular trends show that over an 8-week period the number of cases and deaths are increasing more rapidly in middle and lower HDI states while the curve has flattened in most high HDI states. As the epidemic is spreading covid-19 cases and deaths are more among the lower HDI states and at low SES locations.

Transition of infectious diseases from rich to poor and endemicity of the disease in the latter groups has been known for centuries.^4^ The current covid-19 epidemic is no exception. Covid-19 clinical registries from China and Europe have reported a specific phenotype that is more prone to mortality from this infection-older age, male sex, hypertension, diabetes, obesity, concomitant cardiovascular disease and heart failure-especially in the low SES individuals.^20^ Ethnic minorities are especially susceptible.^21^ Wadhera et al studied population characteristics across the 5 New York boroughs to evaluate differences in covid-19 related outcomes.^15^ It was observed that number of patients with covid-19 who were hospitalized per 100 000 population was highest in the Bronx (634) and lowest in Manhattan (331). The number of deaths related to covid-19 per 100 000 population was also highest in the Bronx (224) and lowest in Manhattan (122). Borough of the Bronx had the highest proportion of ethnic minorities, the most persons living in poverty and the lowest levels of educational attainment as compared to other boroughs of New York.^15^ The UK Office of National Statistics has reported that covid-19 death rates are almost double in most vs. least deprived areas.^22^ The age-standardized mortality rate involving covid-19 in the most deprived areas was 55.1/100,100 compared to 25.3/100,000 in the least deprived areas. London had the highest death rate with high rates in low SES locations of Newham, Brent and Hackney. It has been argued that these minorities are the missing victims of covid-19 infection.^23^ In the present study we have not evaluated state-level social and healthcare characteristics. However, in a previous study it has been reported that state-level HDI correlated strongly with state-level social development index (a composite of 20 social and economic variables).^24^ Indian data also show that more than two-thirds of the patients are concentrated in 13 Indian cities and more than half in the 6 cities mentioned above.^25^ A major study limitation is lack of availability of data from rural India and large central and eastern Indian states.^16^ More studies regarding socioeconomic macro- and micro-level determinants of covid-19 infection are required from India and other low and lower-middle income countries as it has been predicted that covid-19 epidemic shall ultimately reside among the lower socioeconomic stratum in these countries.^23^

A number of studies have predicted the burden of covid-19 infection in India using various modeling techniques, e.g. Bhardwaj, Ghosh et al.^26,27^ These studies focus on extent of the disease and date/s of epidemic ascent and descent. However, modeling techniques of a novel epidemic are not always accurate.^28^ These techniques have been criticized for being insensitive to data quality, intervention strategies and strength of research enterprise.^29^ All these high-quality inputs are missing from most of the forecasting equations and are limitation of the present study also. We have focused on sociological aspects of the disease and show that the epidemic is still in evolution and till the time it matures into low HDI states, it is unlikely to terminate soon (Figure 3). However, although we have used raw data available from a non-profit website,^16^ there are certain differences with the official government data used by others,^26,27^ and the prediction may not be accurate.

The essential urban nature of the disease and rapid spread of the disease in slums pose a challenge to control the epidemic in India and other similar countries. Slums are least prepared for the pandemic of covid-19 as most basic needs such as regular water supply, toilets, waste collection and adequate and secure housing are almost non-existent.^30^ Interventions that have been found useful in China and European countries may be impractical.^31^ Space constraints and overcrowding make physical distancing and self-quarantine impractical in most low SES locations globally and also in India. Corburn et al suggest policies for covid-19 control in urban informal settlements.^30^ These include science-based policies for arresting course of the disease, improving general medical care, provision of economic, social and physical improvements, and focus on urban poor including migrants and slum-communities. Microlevel strategies include institution of informal emergency planning committees in every urban informal settlement, moratorium on evictions, guaranteed payments to the poor, deploying trained community health workers, food assistance, waste management plan and plan for mobility and healthcare.^30^ Focus on using social and behavioral science techniques is essential to harness of benefits of such interventions.^32^ Long term planning is crucial because the course of the epidemic cannot be predicted yet.

Public health strategies that have been successful in various high-income countries of Asia and Europe have focused on widespread testing for covid-19 virus using appropriate technologies, social (physical) distancing, quarantining of the cases using either hospital or home isolation and aggressive and rapid tracing of contacts and their isolation.^30,31^ Sridhar suggests the following eight public health interventions for covid-19 control: large scale testing-tracking-isolation; protection of healthcare workers; monitoring hotspots; watching for imports; clear and honest communication; avoiding going back to ‘normal’; lockdowns useful but crude strategy; and investing in finding alternate strategies.^33^ Testing-tracking-quarantining is the most successful strategy but is expensive and requires immense manpower support.^34^ Development of vaccine and preventive medicine is a work in progress while waiting for herd-immunity can lead to disastrous health consequences. Lockdown has led to both macro-micro-level adverse economic consequences in developed countries as well as in India.^35,36^ Our study shows that the covid-19 epidemic in India is still in the intermediate stage and a judicious strategy involving testing and containment could be the most appropriate strategy. This should be combined with physical distancing and personal protection using standard public health practices. The virus is here to stay.

## Data Availability

All the data are included in the article.

## Competing interests

All authors have completed the ICMJE uniform disclosure form at www.icmje.org/coi_disclosure.pd and declare: no support from any organization for the submitted work; no financial relationships with any organizations that might have an interest in the submitted work in the previous three years; no other relationships or activities that could appear to have influenced the submitted work.

---

**Supplementary Table 1:** Human Development Index (HDI) of various states in India

| State | HDI |
| --- | --- |
| Andhra Pradesh | 0.65 |
| Assam | 0.61 |
| Bihar | 0.57 |
| Chandigarh | 0.77 |
| Delhi | 0.75 |
| Gujarat | 0.67 |
| Haryana | 0.70 |
| Himachal | 0.72 |
| Jammu & Kashmir | 0.68 |
| Jharkhand | 0.59 |
| Karnataka | 0.68 |
| Kerala | 0.78 |
| Madhya Pradesh | 0.60 |
| Maharashtra | 0.69 |
| Orissa | 0.60 |
| Punjab | 0.72 |
| Rajasthan | 0.62 |
| Tamilnadu | 0.70 |
| Telangana | 0.66 |
| Uttar Pradesh | 0.59 |
| West Bengal | 0.64 |

## Notes

### Competing Interest Statement

The authors have declared no competing interest.

### Clinical Trial

Not applicable

### Funding Statement

No funding received.

